# Longitudinal Investigation of Social Isolation, Loneliness, and Health Outcomes Among Refugees in a Low-Resource Setting

**DOI:** 10.1101/2025.11.05.25339624

**Authors:** Gulsah Kurt, Philippa Specker, Belinda Liddell, David Keegan, Randy Nandyatama, Atika Yuanita, Rizka Argadianti Rachmah, Joel Hoffman, Angela Nickerson

**Affiliations:** School of Psychology, University of New South Wales, Sydney, Australia; School of Psychological Sciences, University of Newcastle, New Castle, Australia; HOST International, Sydney, Australia; Excelsia University College, Sydney, Australia; Department of International Relations, Universitas Gadjah Mada, Yogyakarta, Indonesia; SUAKA, Indonesian Civil Society Network for Refugee Rights Protection, Jakarta, Indonesia; School of Medicine, Faculty of Medicine and Health, The University of Sydney, Sydney, Australia

## Abstract

**Background:** Social isolation and loneliness are global public health concerns, adversely impacting mental and physical health. There is an urgent need to expand the evidence base with studies focused on at-risk populations in low-resource contexts. This study aimed to examine the longitudinal association of social isolation and loneliness and health outcomes (probable depression and post-traumatic stress disorder (PTSD), and general health), and to identify individual and social determinants of social isolation and loneliness among refugees in a low-resource context.

**Method:** A longitudinal study involving 1,235 culturally and linguistically diverse refugees was conducted over a two-year period across four time points. Social isolation and loneliness, probable depression and PTSD, and general health were measured using self-report instruments across four time points over two years (six-month intervals). Generalised Linear Mixed Models were used to examine the association between social disconnection and health outcomes over time. Linear regression was used to identify individual, social, and contextual factors associated with social isolation and loneliness.

**Results:** Social isolation and loneliness were associated with increased risk of probable depression, PTSD, and poor health over two years. Younger age, higher education, family separation, traumatic experiences, longer time in the protracted displacement setting, and post-displacement stressors significantly predicted higher social isolation and loneliness at baseline.

**Conclusion:** The present findings indicate a dose-response relationship between social isolation and loneliness and mental health outcomes among refugees in a low-resource setting. Multilevel interventions addressing individual, social, and contextual factors are needed to tackle social disconnection and improve health among refugees.

## Introduction

Social isolation and loneliness - two common forms of social disconnection that capture the objective and subjective dimensions of social relationships - are increasingly recognized as global public health concerns affecting people of all ages (World Health Organization, 2023). They are linked to greater risk for physical and mental health problems such as cardiovascular diseases, immune system deficiencies, depression, suicide, and premature mortality (Leigh-Hunt et al., 2017) - the effects comparable to or even greater than those of well-established risk factors such as smoking and physical inactivity. Social isolation and loneliness also impose substantial economic costs through increased annual health care expenditure (Mihalopoulos et al., 2020). Given these societal and economic costs, countries such as the UK and Australia have introduced national policies (Goldman et al., 2024), and the World Health Organization (WHO) recently launched a Commission on Social Connection (World Health Organization, 2023), to address social isolation and loneliness as neglected social determinants of health and develop evidence-based strategies for prevention and management. Despite this growing policy traction, the existing evidence base remains limited, with studies predominantly cross-sectional in design and conducted in high-income settings (Goldman et al., 2024). There is a pressing need for longitudinal research in low-resource contexts, particularly among at-risk groups such as refugees and asylum seekers.

Refugees and asylum-seekers experience erosion of their existing social networks and face significant barriers in forming new ones following forced displacement (Strang & Quinn, 2021). Ongoing armed conflict and violence often cause profound personal losses, such as the death or disappearance of family members and friends, as well as societal losses, including the destruction of established social institutions and community organizations that once provided formal structure and support for social connection (Silove, 2013; Tay & Silove, 2017). While conflict and war precipitate the erosion of social networks, the challenges refugees face in post-displacement settings often prolong or intensify social disconnection (Miller & Rasmussen, 2017). Refugees and asylum-seekers contend with restrictive policies, financial difficulties, language barriers, discrimination, and interrupted education, all of which constrain opportunities to form new social relationships and develop social ties (Ager & Strang, 2008). In many post-displacement settings, these barriers are further compounded by the absence of integration policies and sustained uncertainty about the future (Kancs & Lecca, 2018; Shahzad et al., 2025). Such conditions push refugees to the margins of society, arguably placing them at heightened risk of prolonged social disconnection.

A recent systematic review on social isolation and loneliness among refugees revealed the consistent link between social disconnection and physical and mental health problems among those resettled in high-income countries (Nguyen et al., 2024). Studies showed that experiencing social isolation and loneliness significantly increases the risk of developing symptoms of common mental health conditions such as post-traumatic stress disorder (PTSD) and depression. Evidence also indicates that social disconnection is associated with poorer physical health outcomes, including chronic pain, headaches, insomnia, and reduced health-related quality of life. Several risk factors, such as female gender, family separation, previous trauma, and adaptation difficulties, have been identified as contributing to increased risk for social isolation and loneliness (Nguyen et al., 2024). Another systematic review examining socio-ecological factors influencing refugee mental health further corroborated the link between social isolation and psychological distress (Fadhlia et al., 2025). While these studies provide important insights into the adversities associated with social isolation and loneliness among refugees and asylum-seekers, they have primarily focused on those resettled in high-income countries. Little is known about social disconnection in low-resource settings, where the majority of the world’s refugees reside (United Nations High Commissioner for Refugees, 2024). This represents an important gap in the evidence base as refugees in these settings usually live in a prolonged transitory state, awaiting resettlement to a third country, and therefore, experience persistent uncertainty and insecurity with limited access to social services(Nickerson et al., 2023). These conditions may further exacerbate social disconnection and its negative impact on health.

The present study aimed to fill this gap by examining the longitudinal association of social disconnection, social isolation and loneliness with mental (depression and PTSD) and general health over a two-year period among refugees in a protracted setting in Indonesia. Furthermore, we sought to identify the individual and socio-contextual determinants of social isolation and loneliness in this sample to highlight potential risk factors. Indonesia is one of the major transit settings in the Western Pacific region, hosting over 12,000 refugees and asylum-seekers (United Nations High Commissioner for Refugees, 2025). As a non-signatory to the 1951 Refugee Convention and its 1967 protocol, asylum-seeking conditions in Indonesia is severely resource-constrained with limited access to healthcare and education and no formal employment opportunities, while awaiting for years, if not decades, for resettlement (Tan, 2016). Thus, the Indonesian context represents a critical transit setting to investigate how sustained social disconnection relates to both mental and physical health over time, offering important insights into the health and social consequences of prolonged displacement.

## Method

### Study Design

The present study used data from a four-wave online cohort study conducted between 2020 and 2022 with refugees living in Indonesia. The first survey was administered between February and October 2020, with subsequent waves conducted at six-month intervals. Participants were recruited via convenience and snowball sampling, including advertisements in refugee services and community-led organizations, and on social media. This recruitment strategy was appropriate given the hard-to-reach nature of the target population. The inclusion criteria were: (1) having arrived in Indonesia as a refugee or asylum seeker in 2013 or later, (2) being aged 18 years or older, and (3) being literate in one of the study languages (Arabic, Farsi, Dari, Somali, or English). These language groups represented the majority of refugees in Indonesia at the beginning of the study(United Nations High Commissioner for Refugees, 2018). Prospective participants provided their consent to participate in the study by filling out the online consent form at the beginning of the survey study. Those who consented proceeded to the rest of the survey, which took around one hour to complete, and participants were compensated with an online grocery voucher of $USD7 (IDR100,000). The study was approved by the UNSW Human Research Ethics Committee (HC190494) and Atma Jaya University, Jakarta (0792/III/LPPM-PM.10.05/07/2019).

### Participants

1235 participants (71.6% male) completed the survey at Time 1 (T1), 961 at Time 2, 772 at Time 3, and 748 at Time 4, with an overall retention rate of 60.57%. The average age was 30.53 years (SD = 9.07), and the average length of time since arriving in Indonesia was 5.11 years (SD = 1.62). The largest language group was Dari or Farsi (39.4%), followed by Arabic (30.3%), English (17.5%), and Somali (12.9%). Table 1 describes the sample’s demographic characteristics.

**Table 1.** Sample Characteristics (N= 1,235) at Time 1.

|  |  |
| --- | --- |
| Gender, female (N, %) | 348 (28.2%) |
| Age (M, SD) | 30.53 (9.07) |
| Language background (N, %) |  |
| Farsi/Dari | 486 (39.4) |
| Arabic | 347 (30.3) |
| English | 216 (17.5) |
| Somali | 159 (12.9) |
| Marital status (N, %) |  |
| Married or in a relationship | 565 (45.9) |
| Not in a relationship | 664 (54.1%) |
| Education level (N, %) |  |
| Little or no formal education | 164 (13.4%) |
| Completed primary school | 372 (30.3%) |
| Completed high school | 425 (34.6%) |
| Completed university | 177 (14.4%) |
| Completed other training | 89 (7.3%) |
| Time in Indonesia in years (M, SD) | 5.11 (1.62) |

Compared to participants who completed only T1, those who completed all time points had been in Indonesia longer (*t*(1230) = 5.759, *p* < .001), had more traumatic experiences (*t*(1010) = 2.131, *p* = 0.033) and reported higher post-displacement stress (*t*(1231) = 3.079, *p* = 0.002), and reported higher social isolation and loneliness (*t*(1200) = 3.199, *p* < 0.001). Participants who completed all time points were also more likely to complete the survey in Farsi (χ*2*(1) = 12.467*, p* < 0.001) and Dari (χ*2*(1) = 9.896*, p* = .002), but less likely to complete the survey in English (χ*2*(1) = 9.797*, p* = 0.002) than Arabic. All-time completers were more likely to report poorer physical health than the T1-only completers (χ*2*(1) = 7.596*, p* = 0.006).

## Measures

### Social isolation and loneliness

Social isolation and loneliness were measured with a single item from the adapted version of the Post-migration Living Difficulties Checklist (Steel et al., 1999) for the Indonesian context at each time point. Participants were asked to rate how much social disconnection (social isolation and/or loneliness) had been a problem for them in the past 12 months on a 5-point Likert Scale (1 = was not a problem, 5 = very serious problem). This single-item measure has been widely used with refugee populations(Nguyen et al., 2024) and globally(Surkalim et al., 2022).

### Mental health outcomes

*Probable depression* was assessed using the Patient Health Questionnaire-8 (PHQ-8) (Kroenke et al., 2009), asking participants to rate how often they have been bothered by each symptom in the past two weeks (e.g., little interest or pleasure doing things and feeling down, depressed, irritable, or hopeless) on a 4-point Likert Scale (0 = no at all, 3 = nearly every day) at each time point. A total score of 10 or more is considered clinically significant depression (Kroenke et al., 2009). Accordingly, we created a dichotomous variable for presence (scores 10 or higher) or absence (scores < 10) of probable depression. *Probable PTSD* was assessed with the adapted version of the Posttraumatic Diagnostic Scale for DSM-IV (PDS) (Foa et al., 1997) with four additional items (negative expectations about oneself/the world, distorted self or other blame, negative emotional states, and reckless behavior) reflecting the revised PTSD symptoms per the DSM-5. A total of 20 items were presented to the participants to rate how often they have been bothered by each over the past month on a 4-point Likert Scale (0 = not at all/only once, 3 = 5 + times a week/almost always). To calculate probable PTSD, the items were considered present if they were rated as 1 (once a week/at least once in a while). Consistent with DSM-5 diagnostic criteria, participants were categorized as meeting the criteria for probable PTSD if they endorsed at least one of the re-experiencing symptoms (Criterion B) and avoidance symptoms (Criterion C), and two of the negative alterations in cognition and mood (Criterion D) and changes in arousal and reactivity symptoms (Criterion E)(Foa et al., 1997) at each time point.

### General health outcome

General health was measured by one global item asking participants to rate their overall health during the past four weeks on a 5-point Likert Scale (1= excellent, 2 = very good, 3 = good, 4 = fair, 5 = poor). The response options were dichotomized as 1 = good (1, 2, and 3) and 0 = poor (4 and 5)(Chen et al., 2019) at each wave. A one-item measure of self-reported health has been widely used and validated in epidemiological studies with the general population(DeSalvo et al., 2006; Macias et al., 2015) and studies with refugees(Chen et al., 2019).

### Potentially traumatic experiences

Experiencing potentially traumatic events was measured using the 19-item scale of the Harvard Trauma Questionnaire (HTQ) (Mollica et al., 1992), on which participants indicated whether they had experienced and/or witnessed each of the listed traumatic events (e.g., imprisonment, torture, or injury) by responding yes (1) or no (0). A total score was calculated by summing the endorsed items.

### Post-displacement related stressors

An adapted version of the Postmigration Living (Steel et al., 1999)Difficulties Checklist with 41 items (excluding the item for social isolation and loneliness) was used to assess the stressors encountered in daily life in the past 12 months in Indonesia. The checklist encompasses items reflecting social, economic, and structural determinants of health and was adapted by the research team for the Indonesian context. A mean score across items was calculated to index the overall level of stressors experienced in the post-displacement context.

### Demographic variables

The demographic information on age, gender, education level, and language was collected as part of the survey package.

### Statistical Analysis

We conducted Generalised Linear Mixed Models (GLMM) using the glmmTMB package in R with a logic link function to investigate whether the change in social isolation predicts the changes in the probability of depression, PTSD, and poor general health over time, after controlling for the key demographic covariates (gender, age, language, and education). The adjusted odds ratios (aOR) and their 95% confidence intervals were calculated to assess the impacts of the observed effects. To address the second objective of the study, we conducted a linear regression analysis in which social isolation at Time 1 was regressed on the individual factors such as gender (female vs. male), age, language groups (dummy coded; comparing Arabic speaking group to English, Farsi, Dari, and Somali speaking groups), education level, (dummy coded; completed tertiary education vs. completed high school and completed tertiary education vs. primary school or no education) and socio-contextual factors such as length of stay in Indonesia, separation from family, exposure to potentially traumatic events, and post-displacement stressors. Multiple imputation using the Multivariate Imputation by Chained Equation (MICE) method was conducted to account for the missing data on the variables in the analyses. We conducted sensitivity analyses on the raw data (without imputation) to assess the robustness of our findings. All the analyses were conducted in R Studio 4.4.1 (R Studio Team, 2020) and the codes and sensitivity analysis results are available in Supplementary Materials. The results from the imputed dataset are presented below.

## Results

The prevalence of probable depression was 54.7% at Time 1, 51.1% at Time 2, 50.2% at Time 3, and 49.7% at Time 4. The prevalence of probable PTSD was 44.9% at Time 1, 42.3% at Time 2, 42.6% at Time 3, and 42.5% at Time 4. The percentages of participants reporting poor overall health were 60.8%, 56.2%, 53.8%, and 52.8% at each respective time point.

### The longitudinal association of social isolation and loneliness with health outcomes

Table 2 presents the results of the models with time and social isolation/loneliness predicting each health outcome. GLMMs showed that social isolation and loneliness were associated with increased risk of probable depression (OR = 2.160, 95% CI 2.025, 2.303) and probable PTSD (OR = 1.780, 95% CI 1.650, 1.921) at any given time point over the 2-year study period. That is, higher levels of social isolation and loneliness almost doubled the risk for probable depression and PTSD at each time point. Higher isolation and loneliness were also associated with a 32% lower likelihood of reporting good health at any given time point over a 2-year period. (OR = 0.676, 95% CI 0.635, 0.721). The main effect of time was significant for depression and general health. Specifically, the odds of probable depression decreased by 11% over time (OR = 0.886, 95% CI: 0.818, 0.960), while the odds of reporting good health decreased by 13% over time (OR = 0.873, 95% CI: 0.812, 0.939). The interaction of time with social isolation and loneliness was not significant for any of the health outcomes, showing that the association of social isolation/loneliness with these did not change over time.

**Table 2.** GLMM Results for the Covariates-Unadjusted Models Predicting Health Outcomes.

| Predictors | Depression<br>OR (95% CI) | p-value | PTSD<br>OR (95% CI) | p-value | General health<br>OR (95% CI) | p-value |
| --- | --- | --- | --- | --- | --- | --- |
| Time | 0.886<br>(0.818–0.960) | 0.004 | 0.939<br>(0.855–1.031) | 0.183 | 0.873<br>(0.812–0.939) | <0.001 |
| Social isolation<br>and loneliness | 2.160<br>(2.025–2.303) | <0.001 | 1.780<br>(1.650-1.921) | <0.001 | 0.676<br>(0.635–0.721) | <0.001 |
| Time × Social<br>isolation/loneliness | 1.048<br>(0.999–1.100) | 0.057 | 1.037<br>(0.982-1.095) | 0.183 | 0.989<br>(0.942–1.039) | 0.662 |

The covariate-adjusted models (Table 3) yielded a significant interaction of time by social isolation, with every unit increase in time, the association between social isolation/loneliness and depression strengthened by 5.1% (aOR= 1.051, 95% CI 1.001-1.103). The remaining results were similar to the initial model.

**Table 3.** GLMM Results for Covariates-Adjusted Models Predicting Health Outcomes.

| Predictors | Depression<br>OR (95% CI) | p-value | PTSD<br>OR (95% CI) | p-value | General health<br>OR (95% CI) | p-value |
| --- | --- | --- | --- | --- | --- | --- |
| Time | 0.888<br>(0.819–0.962) | 0.004 | 0.940<br>(0.856–1.032) | 0.183 | 0.874<br>(0.813–0.939) | <.001 |
| Social isolation and loneliness | 2.092<br>(1.964–2.227) | <.001 | 1.715<br>(1.585–1.857) | <.001 | 0.661<br>(0.620–0.704) | <.001 |
| Time × Social<br>isolation/loneliness | 1.051<br>(1.001–1.103) | 0.044 | 1.042<br>(0.987–1.099) | 0.137 | 0.990<br>(0.943–1.039) | 0.672 |
| Gender (ref = male) | 1.129<br>(0.870–1.466) | 0.353 | 0.917<br>(0.739–1.138) | 0.430 | 0.711<br>(0.563–0.899) | 0.004 |
| Age | 1.000<br>(0.989–1.012) | 0.933 | 0.992<br>(0.980–1.003) | 0.154 | 0.969<br>(0.955–0.982) | <.001 |
| Language<br>(ref = Arabic speaking) |  |  |  |  |  |  |
| English | 1.898<br>(1.392–2.587) | <.001 | 3.243<br>(2.278–4.617) | <.001 | 2.216<br>(1.587–3.094) | <.001 |
| Farsi | 2.609<br>(1.935–3.517) | <.001 | 1.081<br>(0.770–1.519) | 0.647 | 1.089<br>(0.755–1.571) | 0.644 |
| Somali | 1.563<br>(1.098–2.225) | 0.013 | 1.185<br>(0.837–1.676) | 0.335 | 0.824<br>(0.586–1.160) | 0.266 |
| Dari | 2.228<br>(1.655–3.000) | <.001 | 2.362<br>(1.665–3.351) | <.001 | 0.592<br>(0.438–0.800) | <.001 |
| Education level (ref = tertiary<br>education) |  |  |  |  |  |  |
| High school | 0.805<br>(0.588–1.101) | 0.172 | 0.693<br>(0.510–0.941) | 0.019 | 1.008<br>(0.727–1.400) | 0.959 |
| Primary school or no<br>formal education | 1.019<br>(0.718–1.446) | 0.915 | 0.601<br>(0.432–0.836) | 0.002 | 0.669<br>(0.478–0.934) | 0.018 |

### Individual and socio-contextual determinants of social isolation and loneliness

Linear regression analysis (Table 4) identified several significant factors associated with social isolation and loneliness at Time 1. Among individual factors, younger age was associated with higher levels of reported social isolation and loneliness (β = - 0.010, *p* = 0.004). Compared to Arabic-speaking participants, those who completed the survey in English, Farsi, and Dari reported significantly higher levels of social isolation and loneliness (β = 0.369, *p* < 0.001; β = 0.546, *p* < 0.001; and β = 0.286, *p* = 0.005, respectively). Participants with tertiary education reported greater social isolation and loneliness than those with no or primary school education (β = - 0.216, *p* = .035). As for social factors, not having any family members in Indonesia was associated with increased levels of social isolation and loneliness (β = 0.284, *p* < .001). Greater exposure to traumatic experiences (β = 0.027, *p* = 0.022), longer time spent in Indonesia (β = 0.049, *p* = 0.021), and experiencing higher levels of post-displacement stress (β = 1.067, *p* < 0.001) were all associated with a higher risk of social isolation and loneliness.

**Table 4.**
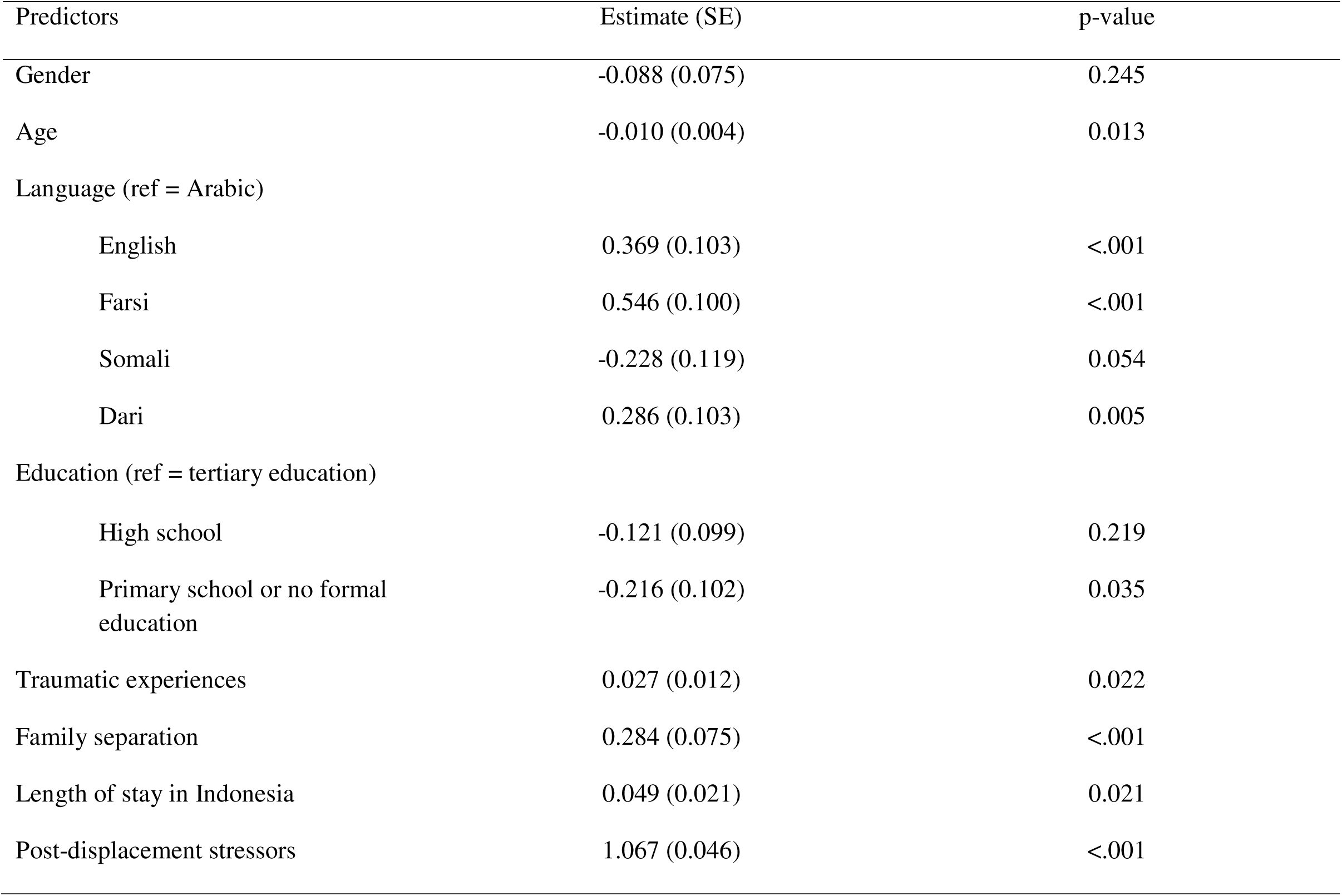
Linear Regression Results for Social Isolation and Loneliness.

| Predictors | Estimate (SE) | p-value |
| --- | --- | --- |
| Gender | -0.088 (0.075) | 0.245 |
| Age | -0.010 (0.004) | 0.013 |
| Language (ref = Arabic) |  |  |
| English | 0.369 (0.103) | <.001 |
| Farsi | 0.546 (0.100) | <.001 |
| Somali | -0.228 (0.119) | 0.054 |
| Dari | 0.286 (0.103) | 0.005 |
| Education (ref = tertiary education) |  |  |
| High school | -0.121 (0.099) | 0.219 |
| Primary school or no formal education | -0.216 (0.102) | 0.035 |
| Traumatic experiences | 0.027 (0.012) | 0.022 |
| Family separation | 0.284 (0.075) | <.001 |
| Length of stay in Indonesia | 0.049 (0.021) | 0.021 |
| Post-displacement stressors | 1.067 (0.046) | <.001 |

## Discussion

The present study is the first to identify individual and social determinants of social isolation and loneliness, and their longitudinal association with both mental health and general health outcomes among refugees in a low-resource setting. Overall, our findings revealed several factors that increase the risk of experiencing social isolation and loneliness, as well as persistent adverse mental and general health outcomes over a two-year period. Given the paucity of research on social isolation and loneliness in low-resource settings and among higher-risk populations such as refugees, this study is uniquely positioned to demonstrate the adversities associated with social isolation and loneliness at the intersection of contextual disadvantages and population vulnerability.

The consistent significant main effect of social isolation and loneliness on mental health outcomes (namely probable depression and PTSD) and general health highlighted that social isolation/loneliness was a robust risk factor for experiencing adverse health outcomes over the two-year study period. Notably, we showed a *dose-response* relationship between reporting a higher level of social isolation/loneliness and probable depression and PTSD. The probability of having depression and PTSD at any given measurement time over the study period was almost doubled by experiencing greater social isolation and loneliness. Similarly, greater social isolation and loneliness were significantly associated with a decreased likelihood of reporting overall good health by approximately one-third of the cohort. While existing studies in the general population and among refugees in high-income settings have shown the association between social isolation and loneliness and various health outcomes(Leigh-Hunt et al., 2017; Nguyen et al., 2024), our findings extend this literature in several key ways. First, this is the first study to provide longitudinal evidence on the adverse health outcomes associated with social isolation and loneliness among refugees in a low-resource, protracted displacement context. Second, our findings highlight that the adverse health risks associated with social isolation and loneliness remain consistently elevated over time across two years. More specifically, a significant interaction effect of time by social isolation and loneliness in the covariate-adjusted model indicated even greater risk for depression over time. Together, these findings underscore that social isolation and loneliness represent a persistent threat to health among refugees in low-resource settings.

Considering the negative health outcomes associated with social isolation and loneliness, it is crucial to identify factors that are linked to a higher risk of these conditions in order to determine effective prevention and intervention strategies. In this regard, our findings on the social and individual determinants of social isolation and loneliness offer important insights into who may be at heightened risk and, therefore, more vulnerable to adverse health outcomes. Some of these determinants align with the cumulative literature on social isolation and loneliness (Barjaková et al., 2023), while others highlight risk factors that are more pronounced in refugee experiences. Specifically, being separated from family, being exposed to a greater number of traumatic events, spending more time in Indonesia, and experiencing more post-displacement stressors, such as prolonged waiting for resettlement, lack of financial assistance, and experiences of discrimination and exclusion, all significantly predicted a higher level of social isolation and loneliness. These findings are particularly important for understanding refugees’ experiences, as they draw our attention to the broader structural and social challenges inherent in forced displacement. The Adaptation and Development after Persecution and Trauma (ADAPT) model (Silove, 2013; Tay & Silove, 2017) provides a useful framework for explaining these findings by highlighting the specific function of social roles, relationships, and networks. According to this model, experiences related to war, conflict, and forced displacement threaten or disrupt the core psychosocial pillars necessary for adaptive psychological and social functioning. These pillars are 1) safety and security, 2) social bonds and networks, 3) justice, 4) social roles and identities, and 5) meaningful life. Of particular relevance here is the breakdown of established social bonds through forced separation from family or the loss of loved ones, and erosion of important social roles and identities due to prolonged waiting, financial problems, and discrimination-factors that can profoundly contribute to social isolation and loneliness(Ali-Naqvi et al., 2023; Kurt et al., 2023; Liddell et al., 2021, 2022). Overall, our findings demonstrate how these experiences pronounced in forced displacement can erode the social fabric of refugees’ everyday lives and increase their vulnerability to social isolation and loneliness.

In addition to these social and contextual determinants, our findings revealed important individual risk factors for experiencing higher levels of social isolation and loneliness. We found that age inversely predicted experiencing social isolation and loneliness, consistent with the recent evidence suggesting that younger individuals – including non-refugee youth - are more likely to experience isolation and loneliness (Barjaková et al., 2023; Shovestul et al., 2020). This may be explained by developmental changes and socio-economic factors that younger individuals face, such as identity formation, limited financial stability, and fewer established social networks(Hawkley et al., 2008). These factors are especially salient in the forced displacement context, where young refugees are separated from their family and social networks, denied basic human rights, and face economic hardship(Silove, 2013). Education was found to be another significant factor associated with social isolation and loneliness in the present study. Contrary to prior research (Barjaková et al., 2023), higher education was associated with greater social isolation and loneliness in this study. While previous studies suggest that individuals with higher education tend to have more access to social and economic resources, enabling them to form larger social networks and relationships that protect against social isolation and loneliness(Hawkley et al., 2008; Hutten et al., 2022), our findings indicate the opposite pattern in the refugee context. This may be because refugees with higher education have more to lose due to forced displacement, such as their social status, economic capital, and existing relationships, and therefore experience more social isolation and loneliness compared to their less educated counterparts. In line with this, a study with refugees in Germany showed that experiencing loss of social status and capital following resettlement was indeed a highly stressful experience, negatively impacting their quality of life and mental health(Costa et al., 2020), which might ultimately lead to social isolation and loneliness.

There are some noteworthy limitations to the present study. Firstly, we used a single-item, combined measure for social isolation and loneliness, which might have impacted the accuracy of estimating these experiences. Although a single-item approach is commonly used in the extant literature and has demonstrated good psychometric properties(Surkalim et al., 2022), it does not allow us to disaggregate the relative contributions of social isolation and loneliness to health outcomes. Despite the efficiency for reducing participant burden and validity of a single-item approach, multi-item and more indirect measures of both are recommended when there is no time constraint(Surkalim et al., 2022). Further, both social isolation and loneliness are detrimental to physical and mental health, but they are distinct, albeit related, constructs. Studies suggest that loneliness is a stronger predictor of mental health, while social isolation is more strongly associated with physical health conditions(Hong et al., 2023; Wang et al., 2023). Future research could examine social isolation and loneliness separately to identify their distinct pathways to health outcomes. Secondly, not all refugee groups in Indonesia were represented in the current study as the sample comprised participants who represented the majority of refugees in the country at the onset of the study (United Nations High Commissioner for Refugees, 2025). Lastly, this was an online, self-administered survey study, which may limit the generalizability of our findings to those with lower general and digital literacy.

### Implications

Despite these limitations, the present study offers several important implications for policy and practice. The current findings contribute directly to the accumulation of evidence base for several priority areas identified by the WHO Commission on Social Connection (World Health Organization, 2025), such as identifying the risk and protective factors and investigating the association between social (dis)connection and health. Our findings regarding several individual and social determinants of social isolation and loneliness provide actionable insights for early identification and intervention of at-risk refugees, especially those who are younger, are more educated and report greater trauma and post-displacement-related stressors, before they develop chronic social isolation and loneliness. These findings also underscore the importance of multi-level interventions that address different aspects of socio-ecologies in which refugees live, from structural policy changes to individual-level interventions. At the policy level, it requires a committed policy effort of the host countries to build the necessary social infrastructure conducive to social connection. Initiatives could include removing legal barriers to social participation, such as restrictions for work and education, and creating opportunities for refugees to engage with local communities in inclusive public spaces such as parks (World Health Organization, 2025). At the community level, social interventions involving the design of participatory group activities, befriending, and physical activity initiatives can effectively increase social engagement within refugee communities and with the local community(Hansen et al., 2024). At the individual level, psychological interventions that target negative cognitions about social interactions and strengthen emotional and social skills may help reduce feelings of loneliness and promote social functioning(Hansen et al., 2024). A randomized-controlled trial with resettled refugees in the USA(Goodkind et al., 2020) further supports the effectiveness of this approach by showing that a multi-level intervention targeted at improving refugees’ competence in navigating the resettlement context, enhancing their access to social services, fostering meaningful social roles, and supporting their relationships with the host community was beneficial in reducing social isolation and psychological distress. This suggests that multi-level interventions may offer a promising pathway to simultaneously enhance social connection and reduce the mental health burden of forced displacement.

## Conclusion

Overall, the present study provides evidence for the longitudinal association between social isolation and loneliness and mental and general health outcomes by highlighting several important risk factors. These findings provide a springboard for future research and practice to investigate the mechanisms through which social isolation and loneliness impact health and to test the effectiveness of multi-level interventions to reduce social isolation and loneliness and improve social connection among refugees in low-resource settings.

## Supplementary material

The supplementary material is available online.

## Data availability

Data is not publicly available due to its sensitive nature. Anonymized data may be shared upon reasonable request to the corresponding author.

## Acknowledgments

We would like to thank refugee community members who took part in the present study for placing trust in us and acknowledge the contributions of our study partners, HOST International, SUAKA, and Universitas Gadjah Mada. We would also like to thank Shraddha Kashyap, Mitra Khakbaz, Diah Tricesaria, Zico Pestalozzi, Yunizar Adiputera, and Shaila Tieken for their invaluable contributions to the study.

## Author contributions

G.K.: Conceptualization, Formal analysis, Writing-Original Draft; P.S: Data Curation, Writing-Review & Editing; B.L, D.K., R.N. & A.Y.: Conceptualization, Methodology, Funding Acquisition, Writing-Review & Editing; J.H: Methodology, Project Administration, Data Curation, Writing-Review & Editing; J.H. & R.A.R: Project Administration, Investigation, Writing-Review & Editing. A.N.: Conceptualization, Methodology, Funding Acquisition, Supervision, Writing-Review & Editing. All authors critically reviewed and approved the final version of the manuscript.

## References

Ager, A., & Strang, A. (2008). Understanding Integration: A Conceptual Framework. Journal of Refugee Studies, 21(2), 166–191. 10.1093/jrs/fen016

Ali-Naqvi, O., Alburak, T. A., Selvan, K., Abdelmeguid, H., & Malvankar-Mehta, M. S. (2023). Exploring the Impact of Family Separation on Refugee Mental Health: A Systematic Review and Meta-narrative Analysis. The Psychiatric Quarterly, 94(1), 61–77. 10.1007/s11126-022-10013-8

Barjaková, M., Garnero, A., & d’Hombres, B. (2023). Risk factors for loneliness: A literature review. Social Science & Medicine, 334, 116163. 10.1016/j.socscimed.2023.116163

Chen, W., Wu, S., Ling, L., & Renzaho, A. M. N. (2019). Impacts of social integration and loneliness on mental health of humanitarian migrants in Australia: Evidence from a longitudinal study. Australian and New Zealand Journal of Public Health, 43(1), 46–55. 10.1111/1753-6405.12856

Costa, D., Biddle, L., Mühling, C., & Bozorgmehr, K. (2020). Subjective social status mobility and mental health of asylum seekers and refugees: Population-based, cross-sectional study in a German federal state. Journal of Migration and Health, 1–2, 100020. 10.1016/j.jmh.2020.100020

DeSalvo, K. B., Bloser, N., Reynolds, K., He, J., & Muntner, P. (2006). Mortality prediction with a single general self-rated health question. Journal of General Internal Medicine, 21(3), 267–275. 10.1111/j.1525-1497.2005.00291.x

Fadhlia, T. N., Doosje, B., & Sauter, D. A. (2025). The Socio-Ecological Factors Associated with Mental Health Problems and Resilience in Refugees: A Systematic Scoping Review. Trauma, Violence & Abuse, 26(3), 598–616. 10.1177/15248380241284594

Foa, E. B., Cashman, L., Jaycox, L., & Perry, K. (1997). The validation of a self-report measure of posttraumatic stress disorder: The Posttraumatic Diagnostic Scale. Psychological Assessment, 9(4), 445–451. 10.1037/1040-3590.9.4.445

Goldman, N., Khanna, D., El Asmar, M. L., Qualter, P., & El-Osta, A. (2024). Addressing loneliness and social isolation in 52 countries: A scoping review of National policies. BMC Public Health, 24(1), 1207. 10.1186/s12889-024-18370-8

Goodkind, J. R., Bybee, D., Hess, J. M., Amer, S., Ndayisenga, M., Greene, R. N., Choe, R., Isakson, B., Baca, B., & Pannah, M. (2020). Randomized Controlled Trial of a Multilevel Intervention to Address Social Determinants of Refugee Mental Health. American Journal of Community Psychology, 65(3–4), 272–289. 10.1002/ajcp.12418

Hansen, T., Nes, R. B., Hynek, K., Nilsen, T. S., Reneflot, A., Stene-Larsen, K., Tornes, R. A., & Bidonde, J. (2024). Tackling social disconnection: An umbrella review of RCT-based interventions targeting social isolation and loneliness. BMC Public Health, 24(1), 1917. 10.1186/s12889-024-19396-8

Hawkley, L. C., Hughes, M. E., Waite, L. J., Masi, C. M., Thisted, R. A., & Cacioppo, J. T. (2008). From Social Structural Factors to Perceptions of Relationship Quality and Loneliness: The Chicago Health, Aging, and Social Relations Study. The Journals of Gerontology: Series B, 63(6), S375–S384. 10.1093/geronb/63.6.S375

Hong, J. H., Nakamura, J. S., Berkman, L. F., Chen, F. S., Shiba, K., Chen, Y., Kim, E. S., & VanderWeele, T. J. (2023). Are loneliness and social isolation equal threats to health and well-being? An outcome-wide longitudinal approach. SSM - Population Health, 23, 101459. 10.1016/j.ssmph.2023.101459

Hutten, E., Jongen, E. M. M., Hajema, K., Ruiter, R. A. C., Hamers, F., & Bos, A. E. R. (2022). Risk factors of loneliness across the life span. Journal of Social and Personal Relationships, 39(5), 1482–1507. 10.1177/02654075211059193

Kancs, d’Artis, & Lecca, P. (2018). Long-term social, economic and fiscal effects of immigration into the EU: The role of the integration policy. The World Economy, 41(10), 2599–2630. 10.1111/twec.12637

Kroenke, K., Strine, T. W., Spitzer, R. L., Williams, J. B. W., Berry, J. T., & Mokdad, A. H. (2009). The PHQ-8 as a measure of current depression in the general population. Journal of Affective Disorders, 114(1–3), 163–173. 10.1016/j.jad.2008.06.026

Kurt, G., Ekhtiari, M., Ventevogel, P., Ersahin, M., Ilkkursun, Z., Akbiyik, N., & Acarturk, C. (2023). Socio-cultural integration of Afghan refugees in Türkiye: The role of traumatic events, post-displacement stressors and mental health. Epidemiology and Psychiatric Sciences, 32, e51. 10.1017/S204579602300063X

Leigh-Hunt, N., Bagguley, D., Bash, K., Turner, V., Turnbull, S., Valtorta, N., & Caan, W. (2017). An overview of systematic reviews on the public health consequences of social isolation and loneliness. Public Health, 152, 157–171. 10.1016/j.puhe.2017.07.035

Liddell, B. J., Batch, N., Hellyer, S., Bulnes Diez, M., Kamte, A., Klassen, C., Wong, J., Byrow, Y., & Nickerson, A. (2022). Understanding the effects of being separated from family on refugees in Australia: A qualitative study. Australian and New Zealand Journal of Public Health, 46(5), 647–653. 10.1111/1753-6405.13232

Liddell, B. J., Byrow, Y., O’Donnell, M., Mau, V., Batch, N., McMahon, T., Bryant, R., & Nickerson, A. (2021). Mechanisms underlying the mental health impact of family separation on resettled refugees. Australian & New Zealand Journal of Psychiatry, 55(7), 699–710. 10.1177/0004867420967427

Macias, C., Gold, P. B., Öngür, D., Cohen, B. M., & Panch, T. (2015). Are Single-Item Global Ratings Useful for Assessing Health Status? Journal of Clinical Psychology in Medical Settings. 10.1007/s10880-015-9436-5

Mihalopoulos, C., Le, L. K.-D., Chatterton, M. L., Bucholc, J., Holt-Lunstad, J., Lim, M. H., & Engel, L. (2020). The economic costs of loneliness: A review of cost-of-illness and economic evaluation studies. Social Psychiatry and Psychiatric Epidemiology, 55(7), 823–836. 10.1007/s00127-019-01733-7

Miller, K. E., & Rasmussen, A. (2017). The mental health of civilians displaced by armed conflict: An ecological model of refugee distress. Epidemiology and Psychiatric Sciences, 26(2), 129–138. 10.1017/S2045796016000172

Mollica, R. F., Caspi-Yavin, Y., Bollini, P., Truong, T., Tor, S., & Lavelle, J. (1992). The Harvard Trauma Questionnaire. Validating a cross-cultural instrument for measuring torture, trauma, and posttraumatic stress disorder in Indochinese refugees. The Journal of Nervous and Mental Disease, 180(2), 111–116.

Nguyen, T. P., Al Asaad, M., Sena, M., & Slewa-Younan, S. (2024). Loneliness and social isolation amongst refugees resettled in high-income countries: A systematic review. Social Science & Medicine (1982), *360*, 117340. 10.1016/j.socscimed.2024.117340

Nickerson, A., Hoffman, J., Keegan, D., Kashyap, S., Argadianti, R., Tricesaria, D., Pestalozzi, Z., Nandyatama, R., Khakbaz, M., Nilasari, N., & Liddell, B. (2023). Intolerance of uncertainty, posttraumatic stress, depression, and fears for the future among displaced refugees. Journal of Anxiety Disorders, 94, 102672. 10.1016/j.janxdis.2023.102672

R Studio Team. (2020). RStudio: Integrated development for R. http://www.rstudio.com/

Shahzad, A., Katona, C., & Glover, N. (2025). The psychological impact of spending a prolonged time awaiting asylum. European Journal of Psychotraumatology, 16(1), 2506189. 10.1080/20008066.2025.2506189

Shovestul, B., Han, J., Germine, L., & Dodell-Feder, D. (2020). Risk factors for loneliness: The high relative importance of age versus other factors. PLOS ONE, 15(2), e0229087. 10.1371/journal.pone.0229087

Silove, D. (2013). The ADAPT model: A conceptual framework for mental health and psychosocial programming in post conflict settings. Intervention Journal of Mental Health and Psychosocial Support in Conflict Affected Areas, 11(3), 237.

Steel, Z., Silove, D., Bird, K., McGorry, P., & Mohan, P. (1999). Pathways from War Trauma to Posttraumatic Stress Symptoms Among Tamil Asylum Seekers, Refugees, and Immigrants. Journal of Traumatic Stress, 12(3), 421–435. 10.1023/A:1024710902534

Strang, A. B., & Quinn, N. (2021). Integration or Isolation? Refugees’ Social Connections and Wellbeing. Journal of Refugee Studies, 34(1), 328–353. 10.1093/jrs/fez040

Surkalim, D. L., Luo, M., Eres, R., Gebel, K., Buskirk, J. van, Bauman, A., & Ding, D. (2022). The prevalence of loneliness across 113 countries: Systematic review and meta-analysis. 10.1136/bmj-2021-067068

Tan, N. F. (2016). The Status of Asylum Seekers and Refugees in Indonesia. International Journal of Refugee Law, 28(3), 365–383. 10.1093/ijrl/eew045

Tay, A. K., & Silove, D. (2017). The ADAPT model: Bridging the gap between psychosocial and individual responses to mass violence and refugee trauma. Epidemiology and Psychiatric Sciences, 26(2), 142–145. 10.1017/S2045796016000925

United Nations High Commissioner for Refugees. (2018). *UNHCR Indonesia Monthly Statistical Report*.

United Nations High Commissioner for Refugees. (2024). Figures at a glance. UNHCR Australia. https://www.unhcr.org/au/who-we-are/figures-glance

United Nations High Commissioner for Refugees. (2025). Indonesia Fact Sheet March 2025. UNHCR Indonesia. https://www.unhcr.org/id/media/indonesia-fact-sheet-march-2025

Wang, F., Gao, Y., Han, Z., Yu, Y., Long, Z., Jiang, X., Wu, Y., Pei, B., Cao, Y., Ye, J., Wang, M., & Zhao, Y. (2023). A systematic review and meta-analysis of 90 cohort studies of social isolation, loneliness and mortality. Nature Human Behaviour, 7(8), 1307–1319. 10.1038/s41562-023-01617-6

World Health Organization. (2023). WHO launches commission to foster social connection. https://www.who.int/news/item/15-11-2023-who-launches-commission-to-foster-social-connection

World Health Organization. (2025). From loneliness to social connection: Charting a path to healthier societies: report of the WHO Commission on Social Connection. Geneva: World Health Organization. https://www.who.int/publications/i/item/978240112360

