## Supplementary Materials for "Longitudinal Investigation of Social Isolation, Loneliness, and Health Outcomes Among Refugees in a Low-Resource Setting"

**S1.** GLMM Results for the Covariates-Unadjusted Models Predicting Health Outcomes.

| Predictors | Depression  OR (95% CI) | p-value | PTSSD  OR (95% CI) | p-value | General health  OR (95% CI) | p-value |
| --- | --- | --- | --- | --- | --- | --- |
| Time | 0.875  (0.805- 0.951) | 0.001 | 0.933  (0.854 - 1.020) | 0.131 | 0.834  (0.768 - 0.906) | <.001 |
| Social isolation and loneliness | 2.291  (2.113- 2.483) | <.001 | 1.874  (1.722 - 2.041) | <.001 | 0.657  (0.608 - 0.710) | <.001 |
| Time × Social isolation/loneliness | 1.036  (0.978- 1.098) | 0.225 | 1.030  (0.969- 1.096) | 0.333 | 0.993  (0.940 - 1.050) | 0.822 |

**S2.** GLMM Results for the Covariates-Adjusted Models Predicting Health Outcomes.

| Predictors | Depression  OR (95% CI) | p-value | PTSSD  OR (95% CI) | p-value | General health  OR (95% CI) | p-value |
| --- | --- | --- | --- | --- | --- | --- |
| Time | 0.865  (0.795–0.941) | < .001 | 0.951  (0.869–1.040) | 0.272 | 0.839  (0.772–0.911) | < .001 |
| Social isolation and loneliness | 2.212  (2.037–2.402) | < .001 | 1.808  (1.657–1.972) | < .001 | 0.640  (0.591–0.694) | < .001 |
| Time × Social isolation/loneliness | 1.034  (0.975–1.097) | 0.260 | 1.036  (0.972–1.103) | 0.276 | 0.992  (0.938–1.049) | 0.771 |
| Gender | 1.220  (0.917–1.621) | 0.171 | 0.862  (0.631–1.177) | 0.349 | 0.632  (0.455–0.878) | 0.006 |
| Age | 1.002  (0.988–1.017) | 0.754 | 0.990  (0.975–1.006) | 0.229 | 0.958  (0.942–0.975) | < .001 |
| Language (ref = Arabic) |  |  |  |  |  |  |
| English | 2.412  (1.639–3.550) | < .001 | 4.708  (3.054–7.257) | < .001 | 2.409  (1.526–3.803) | < .001 |
| Farsi | 3.354  (2.275–4.946) | < .001 | 1.177  (0.780–1.776) | 0.437 | 0.951  (0.618–1.464) | 0.819 |
| Somali | 1.848  (1.173–2.911) | 0.008 | 1.209  (0.706–2.072) | 0.489 | 0.775  (0.468–1.284) | 0.322 |
| Dari | 2.776  (1.899–4.058) | < .001 | 2.901  (1.927–4.368) | < .001 | 0.441  (0.286–0.680) | < .001 |
| Education (ref = tertiary education) |  |  |  |  |  |  |
| High school | 0.807  (0.550–1.184) | 0.272 | 0.670  (0.443–1.013) | 0.057 | 1.023  (0.657–1.594) | 0.918 |
| Primary school or no formal education | 0.976  (0.659–1.448) | 0.905 | 0.538  (0.352–0.823) | 0.004 | 0.648  (0.411–1.021) | 0.061 |

**S3.** Linear Regression Results for Social Isolation and Loneliness.

| Predictors | Estimate (SE) | p-value |
| --- | --- | --- |
| Gender | -0.071 (0.083) | 0.392 |
| Age | -0.010 (0.004) | 0.017 |
| Language (ref = Arabic) |  |  |
| English | 0.318 (0.111) | 0.004 |
| Farsi | 0.541 (0.108) | <.001 |
| Somali | -0.332 (0.142) | 0.019 |
| Dari | 0.184 (0.112) | 0.099 |
| Education (ref = tertiary education) |  |  |
| High school | -0.130 (0.106) | 0.220 |
| Primary school or no formal education | -0.226 (0.110) | 0.040 |
| Traumatic experiences | 0.027 (0.011) | 0.012 |
| Family separation | 0.356 (0.083) | <.001 |
| Length of stay in Indonesia | 0.052 (0.023) | 0.026 |
| Post-displacement stressors | 1.074 (0.048) | <.001 |

**S4. Analytical codes for Data Imputation using MICE in R Studio**

Please note that the refined codes are provided here. The re-use of the codes requires reading your data first in any format (e.g., Excel, SPSS).

library(foreign)

library (mice)

prw_data6$aGenDum1 <- as.factor(prw_data6$aGenDum1)

prw_data6$aLanDum1 <- as.factor(prw_data6$aLanDum1)

prw_data6$aLanDum2 <- as.factor(prw_data6$aLanDum2)

prw_data6$aLanDum3 <- as.factor(prw_data6$aLanDum3)

prw_data6$aLanDum4 <- as.factor(prw_data6$aLanDum4)

prw_data6$aEduDum1 <- as.factor(prw_data6$aEduDum1)

prw_data6$aEduDum2 <- as.factor(prw_data6$aEduDum2)

prw_data6$aFSepD <- as.factor(prw_data6$aFSepD)

prw_data6$healthc <- as.factor(prw_data6$healthc)

prw_data6$depd <- as.factor(prw_data6$depd)

prw_data6$ptsdd <- as.factor(prw_data6$ptsdd)

prw_data6$anxd <- as.factor(prw_data6$anxd)

#Centering time and isolation before the imputation#

prw_data6$time_c <- as.numeric(scale(prw_data6$time, center = TRUE, scale = FALSE))

prw_data6$isolation_c <- as.numeric(scale(prw_data6$isolation, center = TRUE, scale = FALSE))

methods <- make.method(subset_prw_data)

methods["PID_up"] <- ""

methods["time_c"] <- ""

methods["time"] <- ""

\# Specify methods for the variables you want to impute- for instance,

methods["aAge_yrs"] <- ""pmm"

methods["atrauma_exp"] <- ""pmm"

methods["aTime_Indo"] <- ""pmm"

methods["isolation_c"] <- ""pmm"

methods["pmld"] <- ""pmm"

methods["aGenDum1"] <- "logreg”

methods["aFSepD"] <- "logreg”

methods["aLanDum1"] <- “logreg”

methods["aLanDum2"] <- “logreg”

methods["aLanDum3"] <- “logreg”

methods["aLanDum4"] <- “logreg”

methods["aEduDum1"] <- “logreg”

methods["aEduDum2"] <- “logreg”

methods["healthc"] <- “logreg”

methods["depd"] <- “logreg”

methods["ptsdd"] <- “logreg”

\# Create a predictor matrix

predictorMatrix1 <- make.predictorMatrix(subset_prw_data)

\# We use “time” as continuous variable here as predictor here.

predictorMatrix1[, "time"] <- 1

predictorMatrix1["time", ] <- 0 # Do not impute 'time' itself

#We include PID in the final dataset, but NOT as a predictor.

predictorMatrix1[, "PID_up"] <- 0

predictorMatrix1["PID_up", ] <- 0

predictorMatrix1["time_c", ] <- 0

predictorMatrix1[, "time_c"] <- 0

\# Run the imputation

imputed_data_social <- mice(data = subset_data, method = methods, predictorMatrix = predictorMatrix1, m = 10, maxit = 50, seed = 123)

\# Inspect the imputation results

summary(imputed_data_social)

plot(imputed_data_social)

**S4. Analytical codes for GLMM with TBM**

Unadjusted model with time, social isolation and loneliness, time x social isolation and loneliness for health outcomes are given below. The below codes were further adapted for covariate-adjusted analyses.

**## Depression as an outcome##**

depression <- with(imputed_data_social,

glmmTMB(depd ~ time_c + isolation_c +

time_c:isolation_c +

(1 | PID_up),

family = binomial, data = NULL))

\# Pool the model results

pooled_dep <- pool(depression)

\# Get the pooled summary with confidence intervals

pooled_summary_dep <- summary(pooled_dep, conf.int = TRUE)

\# Exponentiate estimates and confidence intervals to get odds ratios

pooled_summary_dep <- transform(pooled_summary_dep,

OR = exp(estimate),

OR_lower = exp(`2.5 %`),

OR_upper = exp(`97.5 %`))

\# View the final table

print(pooled_summary_dep)

**##Health as an outcome##**

healthc <- with(imputed_data_social,

glmmTMB(healthc ~ time_c + isolation_c +

time_c:isolation_c +

(1 | PID_up),

family = binomial, data = NULL))

\# Pool the model results

pooled_healthc<- pool(healthc)

\# Show the pooled summary

summary(pooled_healthc, conf.int = TRUE)

pooled_summary_healthc <- summary(pooled_healthc, conf.int = TRUE) %>%

mutate(OR = exp(estimate),

OR_lower = exp(`2.5 %`),

OR_upper = exp(`97.5 %`))

print(pooled_summary_healthc)

**##PTSD as an outcome##**

ptsdd <- with(imputed_data_social,

glmmTMB(ptsdd ~ time_c + isolation_c +

time_c:isolation_c +

(1 | PID_up),

family = binomial, data = NULL))

\# Pool the model results

pooled_ptsdd<- pool(ptsdd)

\# Get the pooled summary with confidence intervals

pooled_summary_ptsdd <- summary(pooled_ptsdd, conf.int = TRUE)

\# Exponentiate estimates and confidence intervals to get odds ratios

pooled_summary_ptsdd <- transform(pooled_summary_ptsdd,

OR = exp(estimate),

OR_lower = exp(`2.5 %`),

OR_upper = exp(`97.5 %`))

\# View the final table

print(pooled_summary_ptsdd)

**S5. Analytical codes for Linear Regression Analysis at Time 1 to identify risk factors for social isolation and loneliness**

**##T1 SOCIAL ISOLATION RISK FACTORS##**

socialisolation_model_t1 <- with(imputed_data_social,

lm(isolation_c ~ atrauma_exp + aGenDum1 + aAge_yrs + aFSepD +

aTime_Indo + aLanDum1 + aLanDum2 +

aLanDum3 + aLanDum4 +

aEduDum1 + aEduDum2 + pmld,

subset = times == 1))

pooled_iso_model_t1 <- pool(iso_model_t1)

summary(pooled_iso_model_t1)
